# C-reactive protein does not predict future depression onset in adolescents: preliminary findings from a longitudinal study

**DOI:** 10.1101/2023.10.26.23297634

**Authors:** Joshua J. Schwartz, Chloe Roske, Qi Liu, Russel H. Tobe, Benjamin A. Ely, Vilma Gabbay

**Affiliations:** Department of Psychiatry and Behavioral Sciences, Albert Einstein College of Medicine, Bronx, NY; The Nathan S. Kline Institute for Psychiatric Research, Orangeburg, NY

**Keywords:** CRP, Depression, Anxiety, Anhedonia, Inflammation, Adolescent, Longitudinal

## Abstract

**Introduction:** Neuroinflammatory processes have been extensively implicated in the underlying neurobiology of numerous neuropsychiatric disorders. Elevated C-reactive protein (CRP), an indicator of non-specific inflammation commonly utilized in clinical practice, has been associated with depression in adults. In adolescents, our group previously found CRP to be associated with altered neural reward function but not with mood and anxiety symptoms assessed cross-sectionally. We hypothesized that the distinct CRP findings in adolescent vs. adult depression may be due to chronicity, with neuroinflammatory effects on psychiatric disorders gradually accumulating over time. Here, we conducted a longitudinal study to evaluate if CRP levels predicted future onset or progression of depression in adolescents.

**Methods:** Participants were 53 adolescents (ages 14.74 ± 1.92, 35 female), 40 with psychiatric symptoms and 13 healthy controls. At baseline, participants completed semi-structured diagnostic evaluations; dimensional assessments for anxiety, depression, anhedonia, and suicidality severity; and bloodwork to quantify CRP levels. Clinical assessments were repeated at longitudinal follow-up after approximately 1.5 years. Spearman’s correlation between CRP levels and follow-up symptom severity were controlled for BMI, age, sex, and follow-up interval and considered significant at the two-tailed, Bonferroni-adjusted *p* < 0.05 level.

**Results:** After correction for multiple comparisons, no relationships were identified between baseline CRP levels and follow-up symptom severity.

**Conclusion:** CRP levels were not significantly associated with future psychiatric symptoms in adolescents in this preliminary analysis. This may suggest that CRP is not a useful biomarker for adolescent depression and anxiety. However, future longitudinal studies with larger sample sizes and incorporating additional indicators of neuroinflammation are needed.

## Introduction

Adolescent depression is a serious, highly prevalent psychiatric illness associated with numerous negative health outcomes including suicide, the second leading cause of death in this age group (Heron, 2018; Whiteford et al., 2013). Adolescent depression is also a strong predictor of depression in adulthood (Lewinsohn, Rohde, Klein, & Seeley, 1999; Weissman et al., 1999; Wilcox & Anthony, 2004; Wolitzky-Taylor et al., 2014), which is ranked by the Institute for Health Metrics as the third leading cause of disability (IHME, 2018). However, adolescent depression varies considerably in course and severity wherein some depressed youth fully recover, while others will have a more severe and progressive illness into adulthood, despite treatment (Birmaher et al., 2000; Costello, Mustillo, Erkanli, Keeler, & Angold, 2003; Curry et al., 2011; Emslie et al., 2015; Emslie et al., 1998; McCauley et al., 1993). Our group and others have showed that anhedonia and reward dysfunction predict worse outcomes among depressed adolescents (Bennik, Nederhof, Ormel, & Oldehinkel, 2014; Gabbay et al., 2015; Liu et al., 2022; McMakin et al., 2012). However, there are no reliable biological predictors of illness trajectory for adolescent depression at present.

One mechanism postulated to play a role in depression development is chronic inflammation, which has been repeatedly linked to altered reward function in both adolescent and adult cohorts (Bradley et al., 2015; Bradley et al., 2019; Freed et al., 2019; Gabbay, Coffey, et al., 2009; Gabbay, Klein, Alonso, et al., 2009; Gabbay, Klein, Guttman, et al., 2009; Gabbay et al., 2010; Liu, Ely, Schwartz, et al., 2021; Miller, Maletic, & Raison, 2009; Zunszain, Hepgul, & Pariante, 2013). A common proxy for systemic inflammation is C-reactive protein (CRP), an acute phase reactant protein produced by hepatocytes in response to immune challenges (Black, Kushner, & Samols, 2004). CRP can be easily quantified in blood samples and is used clinically an indicator of inflammatory states and chronic disease. Convergent findings over the past two decades indicate that CRP levels are consistently elevated in adults with depression, suggesting that CRP might serve as a useful biomarker for depression in this age group (Baumeister, Akhtar, Ciufolini, Pariante, & Mondelli, 2016; Danese et al., 2008; Danner, Kasl, Abramson, & Vaccarino, 2003; Elovainio et al., 2009; Ford & Erlinger, 2004; Haroon et al., 2018; Orsolini, Pompili, Tempia Valenta, Salvi, & Volpe, 2022). However, studies examining CRP in adolescent depression have reported inconclusive results (Baumeister et al., 2016; Chaiton, O’Loughlin, Karp, & Lambert, 2010; Flouri, Francesconi, Midouhas, & Lewis, 2020; Jha et al., 2020; Khandaker, Pearson, Zammit, Lewis, & Jones, 2014), including a recent study by our group that failed to detect a relationship between blood CRP levels and mood or anxiety symptom severity in a clinically diverse sample of 127 adolescents examined cross-sectionally (Liu, Ely, Simkovic, Alonso, & Gabbay, 2021). Intriguingly, however, we observed a significant association between CRP and neural reward function in a related neuroimaging study of 64 adolescents, suggesting a relationship between systemic inflammation and altered reward processing in adolescents with psychiatric symptoms (Liu et al., 2020). To date, only a few studies have examined whether CRP levels predicted later depression severity, again with conflicting findings (Copeland, Shanahan, Worthman, Angold, & Costello, 2012; Flouri et al., 2020; Moriarity et al., 2019). The disparate results of these longitudinal analyses may be partly due to varied clinical characterization strategies (self-report vs. clinician assessment), participant ages (adult vs. adolescent cohorts), or narrow demographic representation that may limit generalizability (Copeland et al., 2012; Flouri et al., 2020; Moriarity et al., 2019).

Here, we aimed to expand this sparse literature and extend our previous cross-sectional work by examining whether CRP levels would predict future psychiatric symptoms in a longitudinally characterized adolescent sample. We adopted a Research Domain Criteria (RDoC)-inspired approach focusing on continuous dimensional measures of symptomatology and a recruitment strategy designed to capture the full range of symptom severity. As such, our baseline sample consisted of psychotropic-medication-free adolescents with clinically significant psychiatric symptoms, including comorbid and subthreshold diagnoses, as well as healthy controls (HC). We hypothesized that adolescents with higher blood CRP levels at baseline would have more severe psychiatric symptoms at longitudinal follow-up.

## Methods

### Participants

Participants were recruited from the New York metropolitan area through affiliated child and adolescent psychiatry outpatient clinics, physician referrals, and advertisements in the community. Participants consisted of 53 adolescents and young adults (age, M ± SD: 14.74 ± 1.92, range 12-20 years; 35 female). At the time of the initial evaluation, 40 presented with clinically significant psychiatric symptoms (age, M ± SD: 14.83 ± 1.87, range 12-20 years; 27 female) and 13 were healthy control (HC) participants (age, M ± SD: 14.54 ± 2.22, range 12-20 years; 8 female) with no history of mental illness, as determined by clinician-administered diagnostic assessments (see below). All participants were recruited from the 127-subject cohort we reported earlier in our cross-sectional study examining CRP and psychiatric symptoms (Liu, Ely, Simkovic, et al., 2021). We invited back clinical subjects with a primary presentation of mood and/or anxiety symptoms as well as HC; participants with a primary presentation of externalizing disorder symptoms were not included. Clinical follow-up averaged approximately 1.5 years (M ± SD: 19.47 ± 9.43 months, range: 11–52 months). The study was approved by the Institutional Review Board (IRB), and written informed consent was obtained from participants aged 18 and older. Those under the age of 18 provided signed assent, and a parent or legal guardian provided signed informed consent.

### Inclusion and exclusion criteria

#### Inclusion criteria for psychiatric cohort

clinically significant current mood or anxiety symptoms based on clinician evaluation; participants were not required to meet diagnostic criteria for any specific disorder. <u>HC participants</u> had no clinically significant current mood or anxiety symptoms and did not meet criteria for any lifetime psychiatric disorder.

#### Exclusion criteria

1) any physical or neurological conditions; 2) pervasive developmental disorder, current psychosis, or substance use disorders; 3) estimated IQ < 80, as assessed by the Kaufman Brief Intelligence Test (KBIT) (A. S. Kaufman, 1990); 4) self-reported Tanner puberty stage <4; 5) a positive drug toxicology test; 6) a positive pregnancy test in females; 7) psychotropic-medication use in the 1–3 months before baseline visit, depending on drug half-life; 8) any inflammatory illnesses, including the common cold, in the 2 weeks before baseline visit; and 9) anti-inflammatory medication use, including over-the-counter remedies, in the 2 weeks before baseline visit.

### Clinical assessments

All clinical assessments were carried out at both the baseline and follow-up visits.

#### Clinical evaluation procedures

All participants were assessed using the Schedule for Affective Disorders and Schizophrenia for School-Age Children–Present and Lifetime Version (KSADS-PL; J. Kaufman et al., 1997), a semi-structured diagnostic interview. A board-certified child/adolescent psychiatrist or a licensed clinical psychologist trained in administering the KSADS-PL carried out the diagnostic evaluation.

Anhedonia severity was assessed using the self-rated Snaith-Hamilton Pleasure Scale (SHAPS; Snaith et al., 1995), a 14-item scale with a total score range of 14 to 56 constructed to minimize age, sex, and cultural influences. The SHAPS is widely used and has been validated in children and adults (De Berardis et al., 2013; Farabaugh et al., 2015). In addition, temporal components of anhedonia severity were assessed using the Temporal Experience of Pleasure Scale (TEPS; Gard, Gard, Kring, & John, 2006), a self-report questionnaire that quantifies both anticipatory (TEPS-AP, score range 10 to 60) and consummatory (TEPS-CP, score range 8 to 48) anhedonia. Unlike other scales used in this study, higher TEPS scores reflect lower levels of anhedonia. Anhedonia was measured using two rating scales because it is a complex clinical phenomenon.

Overall depression severity was measured by the clinician-rated Children’s Depression Rating Scale–Revised (CDRS-R; Poznanski, Freeman, & Mokros, 1985), which was administered to the participant and also a parent/guardian when the participant was under the age of 18. The CDRS-R has 17 items and a score range of 17 to 113. The self-rated Beck Depression Inventory–Second Edition (BDI; A.T. Beck, Steer, & Brown, 1996), with 21 items and a score range of 0 to 63, was also administered.

Anxiety severity was assessed using the self-reported Multidimensional Anxiety Scale for Children (MASC; March, Parker, Sullivan, Stallings, & Conners, 1997), which has been validated in both clinical and non-clinical population. This scale contains 39 items and a score range of 0 to 117.

Suicidality severity was assessed by the 19-item self-rated Beck Scale for Suicide Ideation (BSSI; A. T. Beck, Kovacs, & Weissman, 1979), which evaluates suicidal thinking and has a total score range of 0 to 38.

### C-reactive protein quantification

At baseline visit, all participants had an overnight fasting CRP level analyzed by LabCorp® via high-sensitivity C-Reactive Protein (hs-CRP) tests, which use immunochemiluminometric assays (ICMA). Since some samples with low CRP levels were only reported as below an upper bound (e.g., CRP < 0.1 mg/L), these were treated as equal to the bounding value (e.g., CRP = 0.1 mg/L) for the purpose of analyses. Specifically, one CRP value reported as “< 1.0 mg/L”, one “< 0.1 mg/L”, three reported as “< 0.2 mg/L”, and 12 reported as “< 0.3 mg/L” were re-coded in this way. To address concerns about quantification accuracy, secondary analyses were performed excluding the subjects with re-coded CRP values (see **Supplementary Materials**).

### Body mass index

Body mass index (BMI) is a standard index of body fat, derived from the mass (kg) and height (m) of a person as the ratio of mass/height^2^. As obesity is associated with inflammation, BMI was calculated for inclusion as a covariate in statistical analyses.

### Statistical analyses for clinical measurements

All statistical analyses for clinical measurements were performed in MATLAB 2018b (The MathWorks, Inc.). As CRP distribution was positively skewed (skewness = 4.7344), nonparametric statistics were used in all the analyses, with significance defined as two-tailed Bonferroni-adjusted *p* < 0.05 (equivalent to uncorrected *p* < 0.0071), accounting for multiple comparisons across the seven clinical assessments (i.e., SHAPS, TEPS-AP, TEPS-CP, CDRS-R, BDI, MASC, BSSI). Spearman’s rank correlation was used to assess associations between CRP levels and the follow-up severity of clinical symptoms in the whole sample, and in the psychiatric subgroup with mood and/or anxiety symptoms excluding HC. Age, sex, BMI, and follow-up interval were controlled for as confounds in the analyses. Secondary analyses were performed excluding these confound variables as well as including baseline symptom severity as an additional confound (see **Supplementary Materials**). Power to detect significant effects was estimated using G*Power v3.1.

## Results

### Demographic and clinical features

Demographic and clinical characteristics of the whole sample (N = 53) during both baseline and follow-up evaluations are presented in **Table 1**. As noted in the **Methods**, all participants were psychotropic-medication-free at recruitment. Eleven participants started psychotropic medications after the baseline visit, of whom five participants remained on medications and six had discontinued medications by the time of the follow-up visit.

**Table 1.**
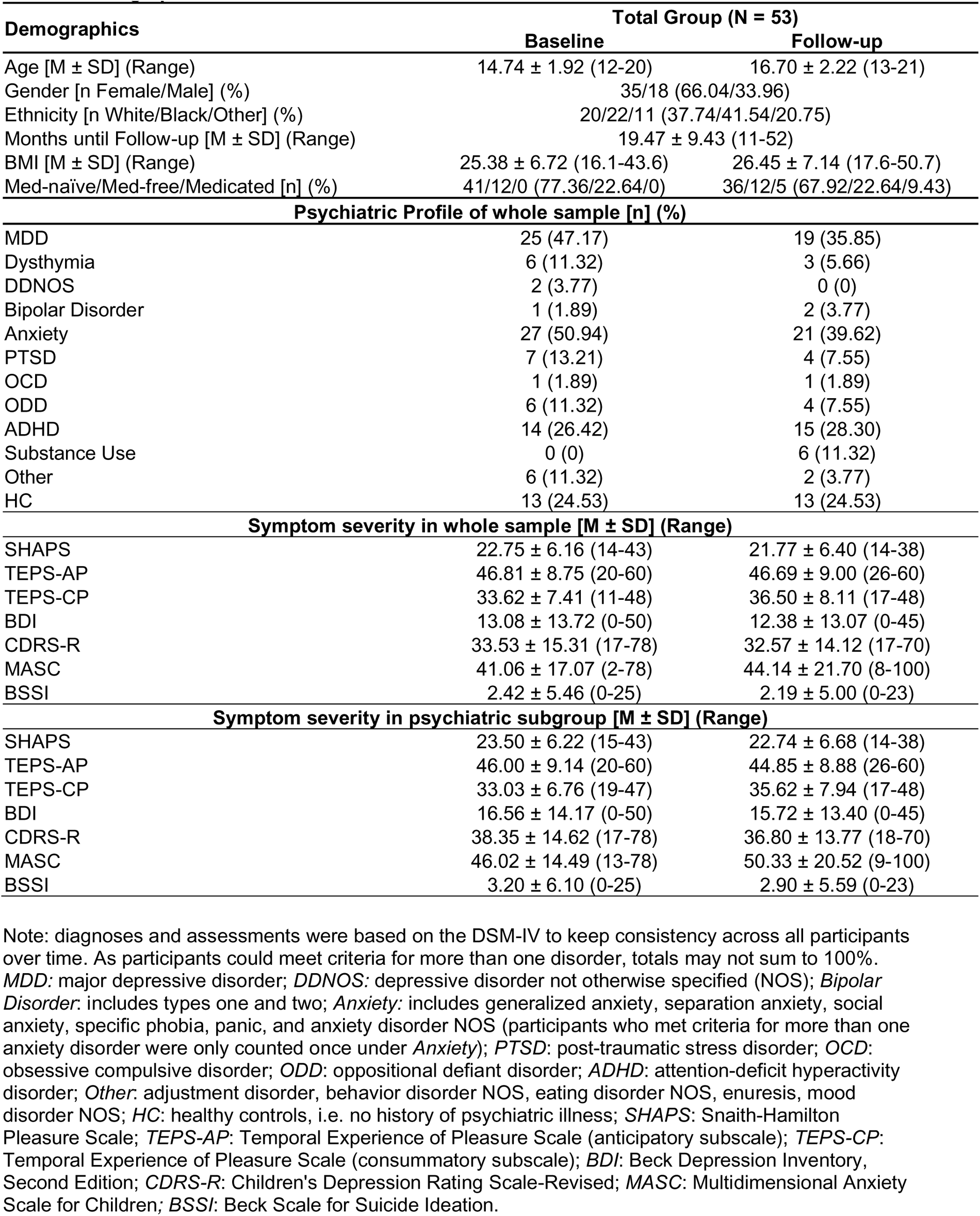
Demographic and Clinical Characteristics.

### Descriptive statistics and power estimation

Blood CRP levels ranged from 0.10-26.20 mg/L (M ± SD = 1.82 ± 3.99) in our whole sample, 0.10-26.0 mg/L (M ± SD = 1.88 ± 4.29) in our psychiatric sample, and 0.20-11.20 mg/L (M ± SD = 1.64 ± 3.02) in the HC sample. There was no significant difference in the CRP levels between the psychiatric sample and the HC sample at baseline (Wilcoxon Rank Sum Test, Z = 0.35, *p* = 0.72). Given the sample of N=53 with two-tailed *α*=0.05 and 1-*β*=0.80, we estimate that our study was powered to detect effects as small as |rho| > 0.37, roughly corresponding to moderate (|rho| > 0.30) or larger correlations.

### Correlation between baseline CRP levels and follow-up symptom severity

Correlation results are represented in **Table 2**.

**Table 2.** Correlation between baseline CRP and follow-up symptom severity.

| **Statistic** | **SHAPS** | **TEPS-AP** | **TEPS-CP** | **CDRS-R** | **BDI** | **MASC** | **BSSI** |
| --- | --- | --- | --- | --- | --- | --- | --- |
| *Whole sample* |  |  |  |  |  |  |  |
| ***N*** | 52 | 52 | 52 | 53 | 52 | 51 | 53 |
| ***rho*** | 0.00 | 0.06 | 0.23 | 0.03 | 0.20 | 0.01 | 0.04 |
| ***p<sub>unc</sub>*** | 0.99 | 0.67 | 0.11 | 0.85 | 0.16 | 0.94 | 0.77 |
| *Psychiatric subgroup* |  |  |  |  |  |  |  |
| ***n*** | 39 | 39 | 39 | 40 | 39 | 39 | 40 |
| ***rho*** | 0.12 | 0.00 | 0.08 | -0.02 | 0.15 | 0.02 | 0.07 |
| ***p<sub>unc</sub>*** | 0.51 | 0.99 | 0.63 | 0.92 | 0.39 | 0.90 | 0.69 |

Analyses controlled for: age, sex, BMI, and follow-up interval.

#### Anhedonia

Anhedonia was measured by SHAPS, TEPS-AP, and TEPS-CP. No significant correlations were identified between baseline CRP levels and follow-up anhedonia measures (SHAPS, TEPS-AP, and TEPS-CP) in the whole sample (all |rho| ≤ 0.23, all *p_unc_* ≥ 0.11) or in the psychiatric subgroup (all |rho| ≤ 0.12, all *p_unc_* ≥ 0.51).

#### Depression

No significant correlations were identified between baseline CRP levels and follow-up depression severity (CDRS-R and BDI) in the whole sample (all |rho| ≤ 0.20, all *p_unc_* ≥ 0.16) or the psychiatric subgroup (all |rho| ≤ 0.15, all *p_unc_* ≥ 0.39).

#### Anxiety

No significant correlations were identified between baseline CRP levels and follow-up anxiety severity (MASC) in the whole sample (rho = 0.01, *p_unc_* = 0.94) or in the psychiatric subgroup (rho = 0.02, all *p_unc_* = 0.90).

#### Suicidality

No significant correlations were identified between baseline CRP levels and follow-up suicidality (BSSI) in the whole sample (rho = 0.04, *p_unc_* = 0.77) or the psychiatric subgroup (rho = 0.07, *p_unc_*= 0.69).

## Secondary analyses

To ensure that potential confounds did not contribute to Type II errors, we also repeated our analyses: a) in unadjusted models to assess the impact of confound variables; b) including baseline symptom severity as an additional covariate to control for initial symptomatology; and c) excluding the 17 subjects whose CRP concentrations were reported as bounded ranges rather than exact numerical values. The results remained the same and no significant associations were found in any of the secondary analyses. See **Supplementary Results** for detailed findings.

## Discussion

This study examined blood CRP levels as a bio-predictor for future psychiatric symptoms in adolescents using association analyses within different clinical cohorts and controlled for potential confounds. Contrary to our hypotheses, we did not detect any significant relationships between CRP levels at baseline and clinical symptomatology at longitudinal follow-up. Previous cross-sectional studies have similarly reported a lack of association between CRP and psychiatric symptoms in pediatric populations (Baumeister et al., 2016; Chaiton et al., 2010; Flouri et al., 2020; Jha et al., 2020; Khandaker et al., 2014; Liu, Ely, Simkovic, et al., 2021). Our results reinforce this negative finding, indicating that CRP is also not predictive of future depression or anxiety symptoms in youth.

Given the well-established association between CRP and depression in adults, the lack of such an association in adolescents may hint at important differences in illness mechanisms across age groups. In particular, older cohorts experience higher rates of medical conditions that drive systemic inflammation; if these conditions also induce parallel neuroinflammatory effects, this could be one reason why CRP is associated with depression in adults but not in youth. While our study evaluated clinical outcomes after a substantial follow-up interval of ∼1.5 years, or approximately 10% of the age of participants, it is possible that underlying levels of systemic inflammation remained too low to induce measurable psychiatric effects within this timeframe.

It also remains possible that inflammatory processes associated with mood symptoms in adolescents may not be as generalized as those in adults. Therefore, more specific markers of inflammation may be needed to probe associations with clinical symptoms in youth. Along those lines, several recent studies have found that levels of interleukin-6 (IL-6), another peripheral inflammatory marker that induces CRP genetic transcription, partially mediated the association between psychosocial stressor exposure in adolescence and later depression (Flouri et al., 2020; Ngwa, Pathak, & Agrawal, 2022). This raises the possibility that neuroinflammation decreases resilience to psychosocial stressors, which may manifest as later clinical symptoms following repeated exposures over time. Despite the apparent lack of association between CRP and clinical symptomatology in adolescence, our team has identified relationships between blood CRP levels and neural reward function in youth, suggesting a mechanistic role for neuroinflammatory processes early in the course of illness (Liu et al., 2020).

Another possibility is that chronic depressive symptoms may induce a systemic inflammatory response. One study found that, although CRP levels in adolescents were not associated with future depression, depressive symptoms displayed an association with increased future CRP levels (Copeland et al., 2012). Another study conducted over five years demonstrated that persistent depressive symptoms were associated with increased future CRP levels, although this association was mediated by smoking (Duivis et al., 2015). Another study documented that stressful life events in conjunction with increased inflammatory response to these events in adolescents predicted increased depression severity at one-year follow-up (Kautz et al., 2020). These studies suggest that the relationship between inflammation and psychiatric symptomatology that has been established by previous literature is not unidirectional and may be multifactorial in nature.

Several limitations should be noted in this study. Foremost is the relatively small sample size of this preliminary investigation, which achieved ∼80% power to detect moderate-to-large associations between CRP and clinical symptomatology. Additionally, while our recruitment strategy aimed to capture a broad range of depression and anxiety symptom severity, it may be reasonable to evaluate relationships with CRP in a more homogeneous clinical sample. However, the wide range of clinical presentations in our sample as well as the inclusion of comorbid and subthreshold conditions allowed for robust dimensional analyses, which were of primary interest in our study and address many of the methodological issues with studying specific categorical diagnoses (Pacheco et al., 2022). Another caveat is our reliance on CRP as a proxy for systemic inflammation; although we believe this was reasonable given past published associations with depressive symptoms, future efforts may benefit from capturing additional and more specific markers of inflammation. Another potential limitation is the wide age range across enrolled adolescents (12–21). However, all participants were Tanner stage ≥ 4 and thus had already achieved the later stages of puberty. Analyses were also adjusted for age as a nuisance covariate to mitigate this concern. Finally, although we included BMI as a confound variable, we did not account for additional factors that might influence inflammatory tone, such as menstrual stage, exercise, diet, stress, and sleep. Thus, while our study advances the understanding of inflammatory processes in adolescents with psychiatric disorders, replication in larger samples while examining other inflammation indices and controlling for additional health and lifestyle factors will be necessary to fully characterize the relationship between inflammation and psychiatric symptomatology in adolescents.

In conclusion, our study recruited adolescents with a diversity of clinical profiles and a wide severity range, including subthreshold diagnoses and HC, and assessed baseline CRP levels with a follow-up clinical evaluation after approximately 1.5 years. The unique sample allowed for robust dimensional analyses across seven distinct measures of overall depression, anxiety, anhedonia, and suicidality. We found no significant relationships between blood CRP levels and future clinical symptom severity. While these results do not preclude the role of the immune system in adolescent mood disorders, they suggest that CRP alone may not be an adequate biomarker to predict future depression symptomatology in youth. Future research should examine larger cohorts over longer time periods and include additional inflammatory indices, such as cytokines, to determine whether immune activation can effectively serve as a biomarker for psychiatric outcomes in adolescents.

## Supporting information

Supplementary Results

## Data Availability

All data produced in the present study are available upon reasonable request to the authors

## Ethical Statement

This study was approved by the Icahn School of Medicine at Mount Sinai Institutional Review Board. The written informed consent was obtained from participants aged 18 and older. Those under the age of 18 provided signed assent, and a parent or legal guardian provided signed informed consent.

## Declaration of competing interest

The authors declare no conflict of interests.

## Role of the funding source

This study was supported by the National Institute of Mental Health (NIMH) grants R01MH101479, R01MH120601, and R21MH121920 to VG (PI).

## Clinical Significance

C-reactive protein (CRP) is a widely used non-specific biomarker for peripheral inflammation and is consistently implicated in adult depression. In adolescents, a significant relationship between blood CRP levels and future psychiatric symptoms has not been detected through group comparison and dimensional analyses. Our negative findings suggest that CRP is not a proper bio-predictor for adolescent depression and anxiety.

## Acknowledgements

The authors would like to acknowledge and thank the participants and their families for their contributions to our study.

## Notes

### Competing Interest Statement

The authors have declared no competing interest.

