## Supplementary Results for "C-reactive protein does not predict future depression onset in adolescents: preliminary findings from a longitudinal study"

### Relationship between baseline CRP and follow-up symptom severity without confounds

The relationship between baseline CRP and follow-up clinical symptom severity in unadjusted models without confounds were examined. CRP levels remained unrelated to any clinical symptoms either in the whole sample (**Supplementary Table 1**, all $|r| \leq 0.18$, all $p_{unc} \geq 0.20$) and in the psychiatric subgroup (all $|r| \leq 0.17$, all $p_{unc} \geq 0.31$).

### Relationship between baseline CRP and follow-up symptom severity controlling for baseline symptom severity

We repeated our analyses controlling for baseline symptom severity as well as age, sex, BMI, and follow-up interval. Just as the findings in the main analyses, relationships between baseline CRP and follow-up clinical symptoms were non-significant among adolescents with detailed CRP levels within the whole sample (**Supplementary Table 2**, all |rho| ≤ 0.20, all $p_{unc} \geq 0.18$) and in the psychiatric subgroup (all |rho| ≤ 0.14, all $p_{unc} \geq 0.42$).

### Relationship between baseline CRP and follow-up symptom severity within the numerical CRP and reported subjects

Lastly, we restricted our analyses to only participants with precisely quantified CRP levels (n=36). Here too, relationships between baseline CRP and follow-up clinical symptoms were non-significant within the whole sample (**Supplementary Table 3**, all |rho| ≤ 0.31, all $p_{unc} \geq 0.09$) and in the psychiatric subgroup (all |rho| ≤ 0.33, all $p_{unc} \geq 0.14$).

**Supplementary Table 1 Correlation between baseline CRP and follow-up symptom severity without confounds**

| Statistic | SHAPS | TEPS-AP | TEPS-CP | CDRS-R | BDI | MASC | BSSI |
|---|---|---|---|---|---|---|---|
| | | | | *Whole sample* | | | |
| ***N*** | 52 | 52 | 52 | 53 | 52 | 51 | 53 |
| ***rho*** | 0.07 | 0.07 | 0.18 | 0.13 | 0.15 | 0.05 | 0.03 |
| $p_{unc}$ | 0.61 | 0.60 | 0.20 | 0.37 | 0.30 | 0.71 | 0.85 |
| | | | | *Psychiatric subgroup* | | | |
| ***n*** | 39 | 39 | 39 | 40 | 39 | 39 | 40 |
| ***rho*** | 0.15 | -0.01 | 0.12 | 0.14 | 0.17 | 0.12 | 0.02 |
| $p_{unc}$ | 0.36 | 0.96 | 0.48 | 0.40 | 0.31 | 0.48 | 0.89 |

Analyses controlled for: no confounds.

**Supplementary Table 2 Correlation between baseline CRP and follow-up symptom severity controlling for baseline symptom severity**

| Statistic | SHAPS | TEPS-AP | TEPS-CP | CDRS-R | BDI | MASC | BSSI |
|---|---|---|---|---|---|---|---|
| | | | | *Whole sample* | | | |
| ***N*** | 52 | 52 | 52 | 53 | 52 | 51 | 53 |
| ***rho*** | 0.01 | 0.01 | 0.17 | -0.10 | 0.20 | -0.14 | -0.08 |
| $p_{unc}$ | 0.95 | 0.95 | 0.25 | 0.50 | 0.18 | 0.36 | 0.57 |
| | | | | *Psychiatric subgroup* | | | |
| ***n*** | 39 | 39 | 39 | 40 | 39 | 39 | 40 |
| ***rho*** | 0.11 | -0.06 | -0.02 | -0.08 | 0.14 | -0.02 | -0.05 |
| $p_{unc}$ | 0.54 | 0.75 | 0.93 | 0.64 | 0.42 | 0.91 | 0.76 |

Analyses controlled for: age, sex, BMI, follow-up interval, and baseline symptom severity.

**Supplementary Table 3 Correlation between baseline CRP and follow-up symptom severity within the numerical CRP reported subjects**

| Statistic | SHAPS | TEPS-AP | TEPS-CP | CDRS-R | BDI | MASC | BSSI |
|---|---|---|---|---|---|---|---|
| | | | | *Whole sample* | | | |
| ***N*** | 35 | 35 | 35 | 36 | 35 | 34 | 36 |
| ***rho*** | 0.17 | 0.04 | 0.14 | 0.13 | 0.31 | 0.24 | 0.18 |
| $p_{unc}$ | 0.37 | 0.81 | 0.47 | 0.46 | 0.09 | 0.21 | 0.33 |
| | | | | *Psychiatric subgroup* | | | |
| ***n*** | 25 | 25 | 25 | 26 | 25 | 25 | 26 |
| ***rho*** | 0.27 | 0.10 | 0.12 | 0.10 | 0.19 | 0.33 | 0.19 |
| $p_{unc}$ | 0.24 | 0.68 | 0.61 | 0.67 | 0.42 | 0.14 | 0.41 |

Analyses controlled for: age, sex, BMI, and follow-up interval.
